# GenECG: A synthetic image-based ECG dataset to augment artificial intelligence-enhanced algorithm development

**DOI:** 10.1101/2023.12.27.23300581

**Authors:** Neil Bodagh, Kyaw Soe Tun, Adam Barton, Malihe Javidi, Darwon Rashid, Rachel Burns, Irum Kotadia, Magda Klis, Ali Gharaviri, Vinush Vigneswaran, Steven Niederer, Mark O’Neill, Miguel O Bernabeu, Steven E Williams

**Affiliations:** King’s College London, London, SE1 7EH, United Kingdom; Guy’s and St Thomas’ NHS Foundation Trust, London, SE1 7EH, United Kingdom; Neurolabs, Edinburgh, EH8 9BT, United Kingdom; University of Edinburgh, Edinburgh, EH8 9YL, United Kingdom

## Abstract

**Background:** Artificial intelligence-enhanced electrocardiogram (AI-ECG) algorithms have primarily been developed using digitised signal data, due to a relative absence of image-based datasets. An image-based ECG dataset incorporating artefacts common to paper-based ECGs, which are typically scanned or photographed into electronic health records, could facilitate clinically useful AI-ECG algorithm development.

This study aimed to create a high-fidelity, synthetic image-based ECG dataset to enable image-based AI-ECG algorithm development.

**Methods:** ECG images were re-created from the PTB-XL database, a publicly available signal-based dataset, and image manipulation techniques were applied to mimic artefacts associated with ECGs in real-world settings. To evaluate the fidelity of the synthetic images, iterative clinical Turing tests were conducted. The ability of currently available algorithms to analyse synthetic ECG images containing artefacts was assessed.

**Results:** Synthetic images were created from all PTB-XL ECGs leading to the creation of GenECG, an image-based dataset containing 21,799 ECGs with artefacts encountered in routine clinical care paired with artefact-free images. Clinical Turing tests confirmed the realism of the images: expert observer accuracy of discrimination between real-world and synthetic ECGs fell from 63.9% (95% CI 58.0%-69.8%) to 53.3% (95% CI: 48.6%-58.1%) over three rounds of testing, indicating that observers could not distinguish between synthetic and real ECGs. The performance of pre-existing image-based algorithms on synthetic (AUC 0.592, 95% CI 0.421-0.763) and real-world (AUC 0.647, 95% CI 0.520-0.774) ECG images containing artefact was limited. Algorithm fine-tuning with GenECG data led to an improvement in classification accuracy on real-world ECG images (AUC 0.821, 95% CI 0.730-0.913) demonstrating the potential for synthetic data to augment image-based AI-ECG algorithm development.

**Conclusions:** GenECG is the first synthetic image-based ECG dataset to pass a clinical Turing test. The dataset will enable image-based AI-ECG algorithm development, ensuring utility in low resource areas, pre-hospital settings and hospital environments where signal data are unavailable.

**What is already known on the subject?:**

- Artificial intelligence-enhanced ECG (AI-ECG) analysis presents a significant opportunity to improve the care of patients with cardiovascular disease.
- Most AI-ECG algorithms have been developed using ECG signal data, limiting their ability to analyse paper-based ECGs which are still prevalent in various hospital and non-hospital settings.

**What this study adds:**

- This study presents GenECG, a high-fidelity, synthetic dataset comprising 21,799 ECG images paired with artefact-free images and ECG signal data.
- GenECG is the first publicly available synthetic, image-based ECG dataset to pass a clinical Turing test.
- The performance of image-based AI-ECG algorithms improved through fine-tuning with GenECG data demonstrating the potential for synthetic data to augment AI-ECG research.

**How this study might affect research, practice or policy:**

- GenECG will facilitate the development of image-based AI-ECG algorithms, promising to expand the application of AI-ECG to traditional hospital settings, reliant on paper-based ECGs, and non-hospital environments such as remote healthcare areas or pre-hospital settings.

## Introduction

The use of synthetic data in healthcare can facilitate the development of high-fidelity, fully anonymised patient datasets on a previously unachievable scale.[1] Synthetic patient data can be used as training data to develop artificial intelligence-enhanced (AI) algorithms offering the potential to revolutionise the scope and utility of AI within healthcare settings. Several studies have already highlighted the potential benefits that AI algorithms may offer when applied to the ECG.[2] For example, AI-ECG can detect electrolyte imbalances,[3] identify left ventricular systolic dysfunction[4] and predict risks of paroxysmal arrhythmia[5] and all-cause mortality.[6] The ability of AI-ECG to facilitate automated ECG interpretation and detect patterns imperceptible to human observers[2] presents a significant opportunity to improve the care of patients with cardiovascular disease.

However, despite the potential uses for AI-ECG, current algorithms are primarily limited to analyses of digitised ECG signals rather than ECG images. This is reflective of the composition of currently available public ECG datasets. Whilst multiple signal-based datasets exist,[7–9] the availability of image-based ECG data is limited. To our knowledge, only one publicly available image-based ECG dataset exists.[10] However, this dataset lacks the artefacts encountered with real-world paper-based ECGs and is substantially smaller than most digital ECG datasets. Nevertheless, numerous healthcare environments continue to rely on printed or scanned ECG images.[11,12] Given the ongoing, widespread use of paper-based ECGs, a disconnect therefore exists between commonly available data types and the AI algorithms designed for their analysis. The creation of a dataset comprising ECG images could enable the development of image-based AI algorithms for use in scenarios where ECG signal data is unavailable. Such a dataset should capture the full range of diversity in paper-based ECGs incorporating artefacts common to clinical practice.

The aims of this study were to 1) create a publicly available image-based ECG dataset of labelled images containing artefacts typically encountered in routine clinical care; 2) demonstrate the fidelity of the synthetic ECGs by testing the discrimination of synthetic from real-world images by healthcare professionals; and 3) evaluate whether the performance of pre-existing image-based AI algorithms could be improved through fine-tuning on synthetic data.

## Methods

### PTB-XL Dataset

Input ECG signals were provided by the PTB-XL database which contains signal data representing 21,799 clinical ECGs from 18,869 patients.[7] PTB-XL ECGs are configured as 12 channel binary files with a resolution of 1μv / LSB at 500Hz (each sample is 0.002 sec). Annotated by two cardiologists, there are 71 different ECG statements within the dataset. The statements cover form, rhythm, and diagnostic labels in a machine-readable form. The diagnostic labels are organised into five superclasses and 24 subclasses as described in [7] (Supplementary Table S1, Figure S1).

### ECG recreation

For each PTB-XL ECG, an image was created according to recommendations outlined in the ‘AHA/ACCF/HRS Recommendations for the Standardisation and Interpretation of the Electrocardiogram’ document,[13] comprising a continuous ten second recording divided into three rows and four columns consisting of 2.5 seconds of data for each lead where column one represents leads I, II and II; column two represents aVR, aVL and aVF; column three represents V1, V2 and V3 and column four represents V4, V5 and V6. An additional rhythm strip containing 10 seconds of data (lead II) was included for rhythm analysis.

The Blender (Blender Foundation, Amsterdam, Netherlands) software platform was used to create synthetic ECG images using custom code (developed by AB). ECG images were recreated by sampling 2.5s epochs for each lead, positioned according to AHA/ACCF/HRS recommendations[13] and with lead markers, lead labels and calibration scales added. Resulting waveform traces were superimposed onto a paper grid (with a resolution set to 150Hz (25mm/s) horizontally and 10.0mm/mV vertically) leading to the generation of a single layout for each PTB-XL ECG. The resolution of the waveform image was set to 5 pixels/mm with a final image output size of 1397 x 1029 pixels for a 10 second trace.

To validate the accuracy of initial ECG images created from signal data, ECG files representing sine waves of known amplitude (0.5mV) and frequency (1.25Hz) were created using the WFDB Toolbox for MATLAB/Octave.[14] A total of 12 test ECGs were created, consisting of ECGs with the sine wave at a single ECG lead location with all other leads set to a constant electrical potential of 0mV. These files were converted into ECG images using the same code used for ECG recreation. All validation ECGs were inspected to confirm the correct location of the ECG leads. For each lead of each ECG containing a sine wave, the amplitude and cycle length (frequency) were measured by an observer (NB) blinded to the original amplitude and frequency. Spearman’s correlation coefficient was used to examine the correlation between measured and actual sine wave frequency and amplitude.

### Creation of synthetic ECG images

To add realistic artefacts to ECG images (i.e. to make it appear as though images had been photographed), ECGs were passed to a second render which placed each image trace on a 3D model comprising a paper sheet positioned in a synthetically developed workspace. In total, 352 unique geometric variations were created from eight paper sheet variations, eleven workspaces and four synthetic workspace orientations. The Blender platform’s bpy module was used to create an automated Python script for ECG image generation. For each ECG, a mesh and synthetic workspace were randomly selected, and the location and rotation of the ECG paper sheet, camera, and light sources were randomly adjusted. To mimic the imperfections associated with photographed ECGs, varying degrees of stucci noise were applied.[15] This technique, which simulates the appearance of stucco (a wall structure containing holes and bumps), was chosen following a review of the artefacts encountered with real-world ECG photographs by a senior 3D technical artist (AB). For each image, the size and turbulence of the noise were randomly selected to introduce varying degrees of texture distortion.

### Clinical Turing Tests

A series of visual Turing tests were designed and conducted to assess the fidelity of synthetic ECG images via an online survey (Qualtrics, Provo, UT). In all rounds of Turing tests, healthcare professionals were provided with a series of 60 images comprising 30 synthetically created ECGs and 30 photographs of real-world ECGs. ECG images were redacted in areas where text may appear. Images were displayed one-by-one to participants and shown in uniform order. Participants were asked to select whether they thought the images were real or synthetic and, in the second and third rounds, to rate their confidence using a five-point Likert scale (Supplementary Figure S2). At the end of each survey, healthcare professionals were asked to provide qualitative feedback through a series of open questions. Feedback was summarised and used to iteratively improve the dataset’s fidelity. All readers decided whether each image was real or synthetic without any time limit and no prior knowledge regarding the number of real or synthetic images. To avoid bias, healthcare professionals were only allowed to complete one round of clinical Turing tests.

For each round of Turing Tests, we measured the Accuracy (overall proportion of ECGs correctly identified as ‘real-world’ or ‘synthetic’), True Recognition Rate (proportion of real-world ECGs identified correctly) and False Recognition Rate (proportion of synthetic ECGs identified correctly) using adapted terminology from previous Turing tests.[16,17] The Fleiss-Kappa score was calculated to evaluate the degree of inter-observer agreement. For the second and third rounds of clinical Turing tests, confidence Likert scale scores were converted to a signed ordinal scale for area under the curve-receiver operating characteristic (AUC-ROC) score analysis. The data were analysed using SPSS version 29 (IBM Corp., Amonk, NY).

### Assessment of pre-existing image-based algorithms

To examine the performance of currently available image-based algorithms on the GenECG dataset, synthetic images were inputted into two image-based AI-ECG algorithms.[11,18]

The first image-based algorithm tested was ECG-Dx© (https://www.cards-lab.org/ecgdx), an automated diagnostic algorithm capable of detecting six diagnoses (atrial fibrillation, sinus tachycardia, sinus bradycardia, left bundle branch block, right bundle branch block and first-degree atrioventricular block). We searched the PTB-XL database for these diagnoses and randomly selected 75 abnormal ECGs. Images with and without image degradation techniques were inputted into the web-based platform. The corresponding classifiers were compared with labels assigned from the PTB-XL dataset.

The second image-based algorithm examined was developed by Bridge *et al.,* to distinguish ‘normal’ from ‘abnormal’ ECGs, and this algorithm has demonstrated good performance on scanned ECG printouts.[18] This model was originally developed using 1172 ECGs and built on InceptionV3,[19] a pre-trained convolutional neural network, with extra layers added to improve performance and prevent overfitting. Due to unavailability of the original model weights and ECG dataset, we trained an identical model using 1682 images from the “ECG images dataset of Cardiac and COVID-19 patients,” an open-access, artefact-free dataset.[10] The dataset contains five distinct categories: COVID-19 (n=250), myocardial infarction (n=74), abnormal heart beat (n=546), history of myocardial infarction (n=203) and normal person ECG images (n=859). Given the anticipated challenges in distinguishing normal versus abnormal ECGs in COVID-19 patients, we excluded images from the COVID-19 category. The remaining 1682 images were defined as normal (n=859) or abnormal (n=823), and randomly split into train (n=1082), validation (n=200) and test datasets (n=400).

To assess the algorithm’s ability to analyse synthetic data, we created 215 synthetic ECG images (119 abnormal, 96 normal) containing image degradation techniques from the PTB-XL dataset. The images were randomly split into train (n=150), validation (n=22) and test (n=43) images. The trained model was applied to evaluate its performance on the 43 test synthetic ECG images. Following the initial results, the model had low efficiency on synthetic GenECG images since it was not exposed to images that resembled our images during training. Therefore, the model was fine-tuned on 215 synthetic images using weights from the model trained on the open-access dataset as initial weights. The additional layers of the Bridge *et al.,* model were adjusted accordingly.

To assess the generalisation power of the synthetic-model and to ensure that the model did not over-fit during fine-tuning, we re-evaluated the trained synthetic model over the open-access test dataset (n=400). Additionally, we evaluated the performance of both the initially developed model and the fine-tuned model on 79 real-world ECG images obtained using methodology described by Sangha *et al.,*[11]. Images which would have been defined as either normal (n=24) or abnormal by the Bridge *et al.,* algorithm (five abnormal ECGs for each label of interest: sinus arrhythmia, atrial fibrillation, atrial flutter, premature atrial contraction, premature ventricular contraction, atrioventricular block, ventricular tachycardia, supraventricular tachycardia, Wolff-Parkinson-White syndrome, paced rhythm, junctional rhythm)[18] were obtained from both the life in the fast lane website (https://litfl.com/ecg-library/) and Google searches. The images contained artefacts typically encountered in routine clinical care.

ECG images were pre-processed by cropping the region of interest using the rembg Python library (https://pypi.org/project/rembg/2.0.28/) to remove the background of each image. Images were then resized using the Lanczos method to ensure uniform input into the image-based model.[20] Subsequently, AUC-ROC analysis was performed to evaluate model performance.

## Results

### ECG recreation

Synthetic ECG images were created for all 21,799 PTB-XL ECGs (Dataset A: ECGs without artefact) (Figure 1A, B). Technical validation confirmed that all ECG leads were plotted in the correct location. There was a perfect correlation between measured and actual sine wave amplitude (R=1.0, P<0.001) and frequency (R=1.0, P<0.001) for each ECG lead.

**Figure 1.**
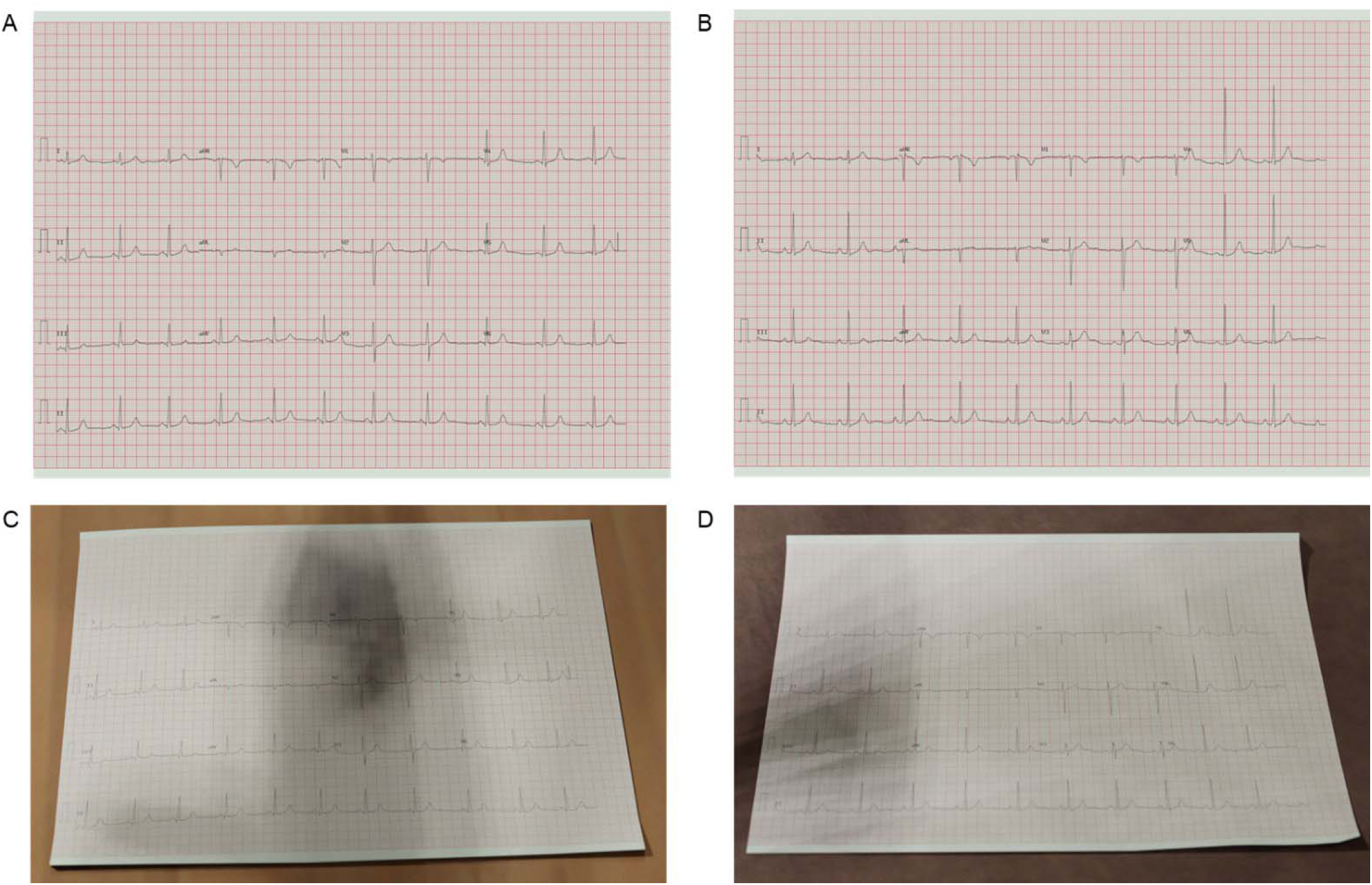
Synthetic ECG images recreated from the PTB-XL dataset. Panels A and B represent artefact-free images. Panels C and D represent the same images following the application of image degradation techniques to make it appear as though the images have been photographed. Panels A and Panels C have been recreated from 00074_hr_1R.dat. Panels B and D have been recreated from 00067_hr_1R.dat.

### Clinical validation

Image degradation techniques were applied to a randomly selected subset of synthetic ECGs for clinical validation. The results of the initial two rounds of Turing Tests indicated that healthcare professionals were able to distinguish real-world ECGs from synthetic images (round one accuracy 63.9% (95% CI 58.0%-69.8%), round two accuracy 59.8% (95% CI 55.9%-63.7%)) (Supplementary Table S2). Qualitative feedback (summarised in Figure 2) was used to iteratively improve the fidelity of the ECG images (Figure 1C, D). In the third round of Turing tests, the accuracy, true recognition rate and false recognition rate were 53.3% (95% CI 48.6%-58.1%), 53.0% (95% CI 48.7%-57.2%) and 53.7% (95% CI 47.4%-60.0%) respectively (Supplementary Table S2). The Fleiss-Kappa score of 0.049 (95% CI 0.007-0.092) indicated a high degree of inter-observer variability. The AUC-ROC curve score was 0.480 (95% CI 0.432-0.529) indicating that level of confidence was not an accurate predictor of an observer’s ability to distinguish real-world ECG from synthetic images (Figure 3).

**Figure 2.**
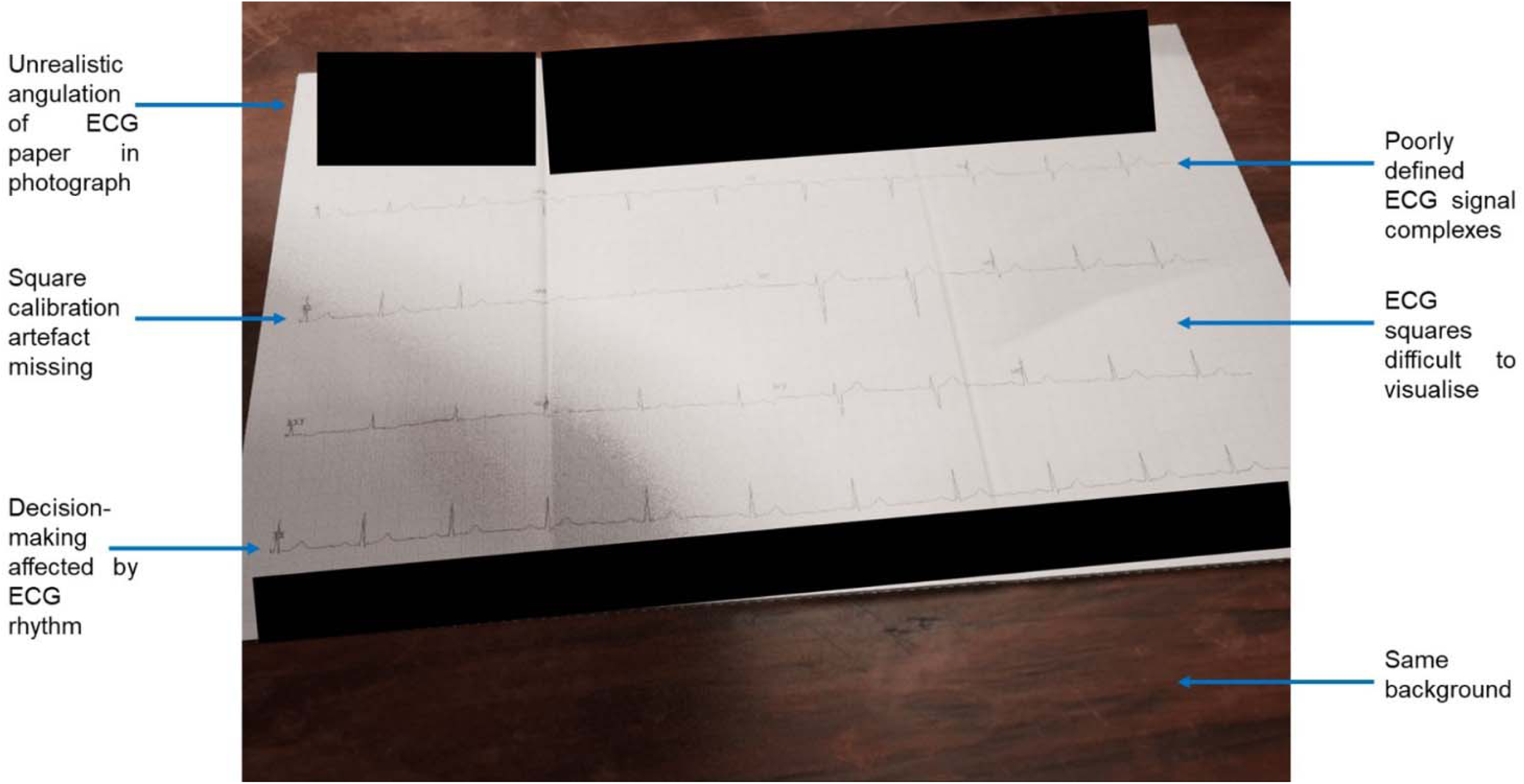
Example of a synthetic ECG used in the initial Turing test with a summary of the qualitative feedback provided by healthcare professionals.

**Figure 3.**
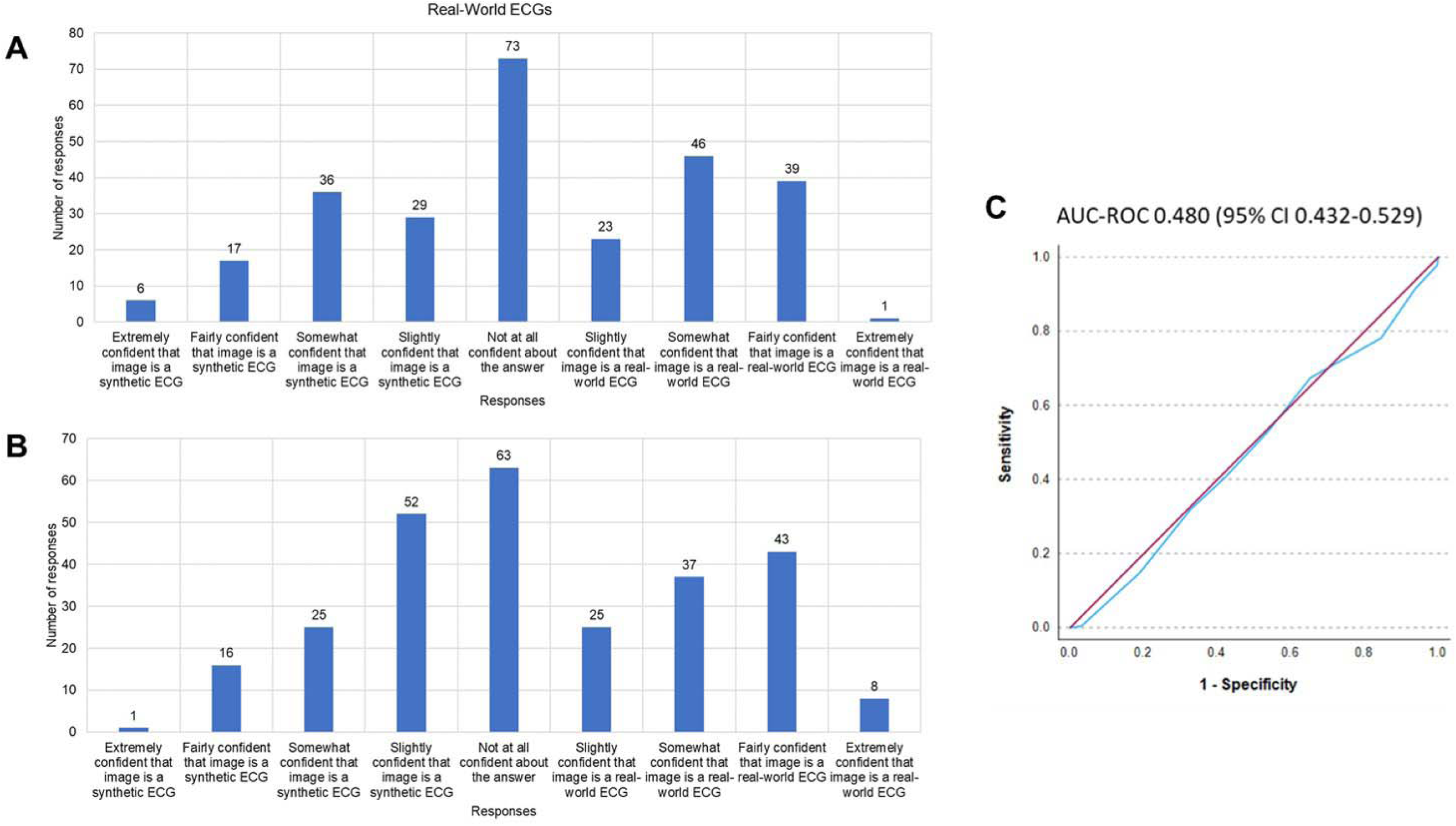
Confidence levels in ECG classification for Turing Test round three. Panels (A) and (B) represent the number of responses for each option for (A) real-world ECGs and (B) synthetically created ECGs. Panel C shows an area under the curve-receiver operating characteristic (AUC-ROC) curve examining the impact of confidence level on ability to correctly identify an ECG image as ‘real-world’ or ‘synthetically created.’

Upon completion of the ‘Turing Tests’, the artefact generation algorithm was deemed capable of recreating life-like ECGs from the PTB-XL database. A dataset of 21,799 images containing PTB-XL ECGs with the incorporation of artefacts common to photographed images was created (Dataset B: ECGs with artefact) (Figure 1C, D; Supplementary Figure S3).

### Assessment on pre-existing image-based algorithms

We assessed the performance of 75 abnormal ECGs both with and without artefact on the ECG-Dx© image-based algorithm.[11] The algorithm was able to correctly identify abnormal diagnoses for 51/75 (68%) artefact-free images but only 29/75 (39%) images containing artefact (Figure 4).

**Figure 4.**
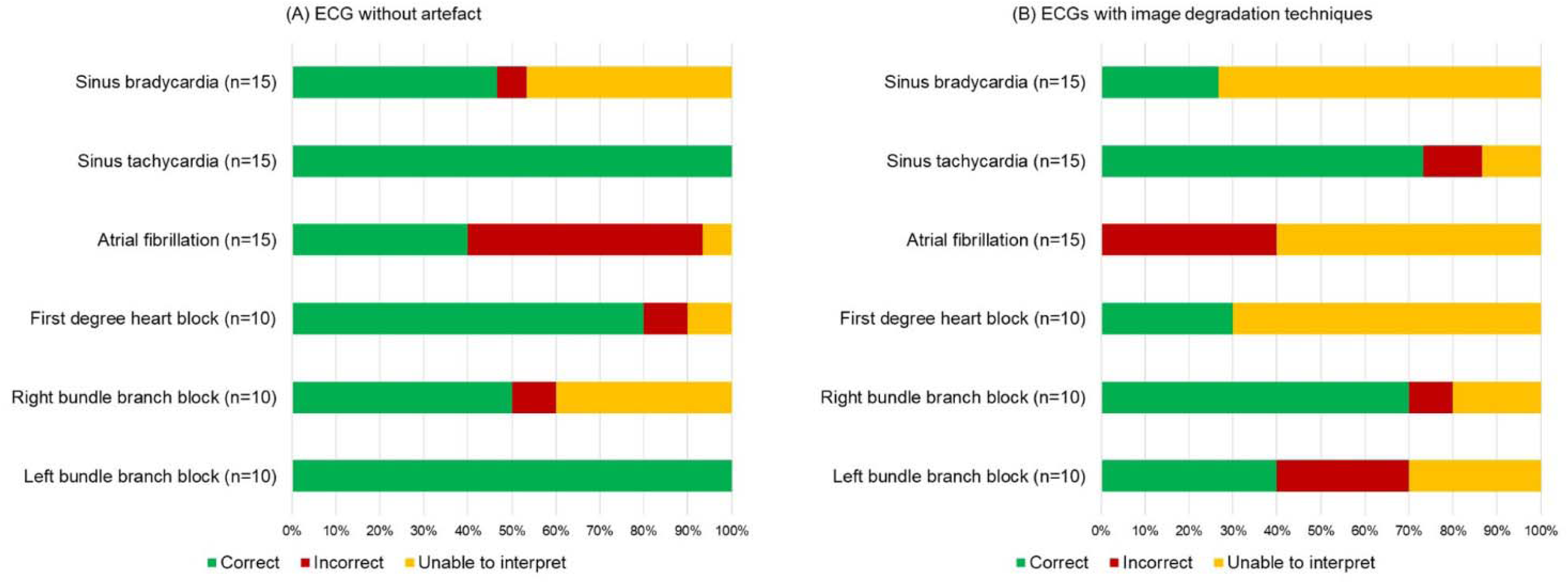
Performance of ECG-Dx**©** image-based algorithm on abnormal images obtained from the PTB-XL dataset. Diagnoses along the Y axis represent PTB-XL labelled diagnoses. The label “Unable to interpret” corresponds to an output of “None” from the image-based algorithm.

Using the same architecture as an image-based model developed by Bridge *et al.,*[18] we trained a synthetic model with artefact-free images from an open access real-world dataset. The synthetic model initially achieved an AUC score of 0.956 (95% CI 0.936-0.977) on 400 artefact-free images (Figure 5A). The model was less able to distinguish normal from abnormal on 43 GenECG images (AUC score 0.592, 95% CI 0.421-0.763) (Figure 5B). Fine-tuning of the model led to a substantial improvement in image classification on the same 43 images (AUC score 0.945, 95% CI 0.876-1.000) (Figure 5C). The fine-tuned model subsequently achieved an AUC score of 0.896 (95% CI 0.864-0.928) on 400 real-world ECGs demonstrating that the model did not over-fit during fine-tuning (Figure 5D). In addition, we assessed the performance of both the initially developed synthetic and the fine-tuned model on 79 real-world ECG images containing artefact (Figure 5E). The initially developed synthetic model achieved an AUC of 0.647 (95% CI 0.520-0.774) whilst the fine-tuned model achieved an AUC of 0.821 (95% CI 0.730-0.913) demonstrating an improvement in performance following model fine-tuning on GenECG data.

**Figure 5.**
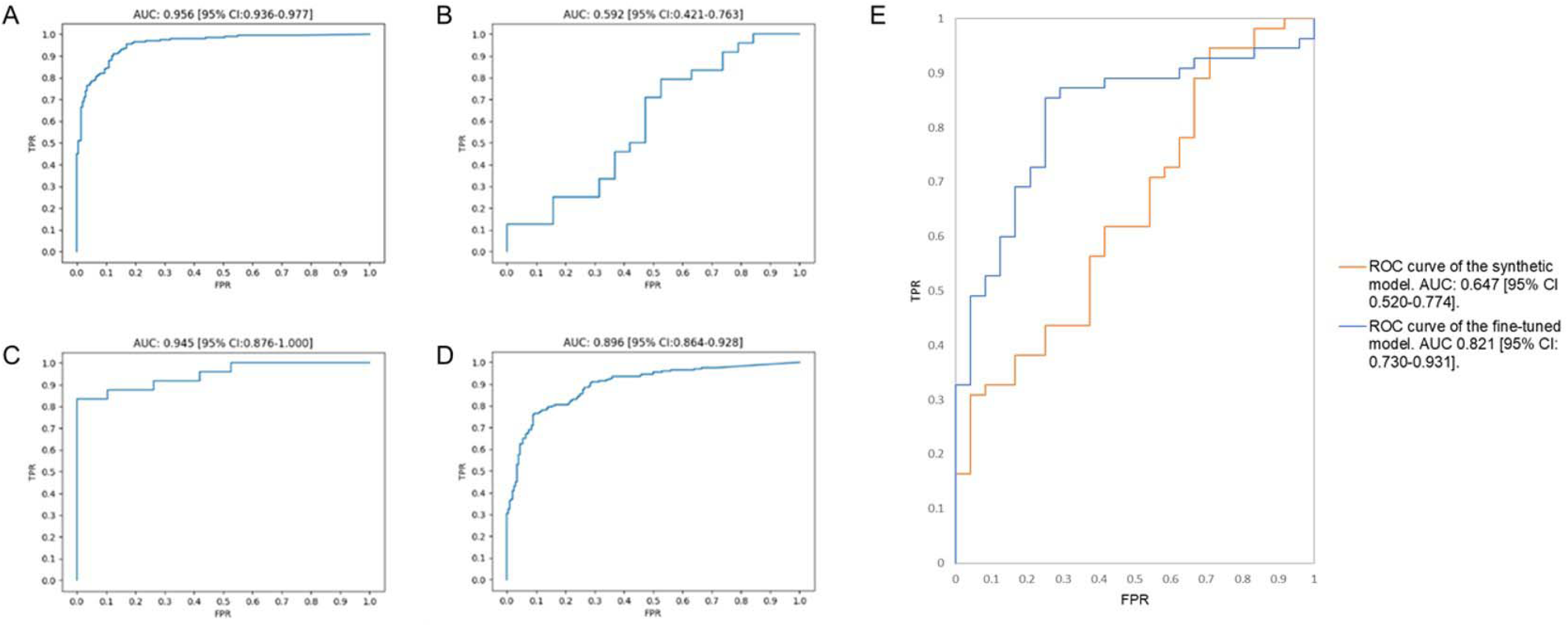
Area under the curve-receiver operating characteristic score curves for ECG images on the algorithm initially developed by Bridge et al., [18]. (A) represents the performance of the model on 400 test images obtained from an artefact-free real-world image-based dataset [10], (B) demonstrates the performance of the trained model on 43 test synthetic ECG images containing artefact, (C) demonstrates the performance of the trained model on the same synthetic ECG images following fine-tuning, (D) shows the performance of the fine-tuned model on the 400 test images from the artefact-free real-world image-based dataset, (E) represents the performance of both the initially developed synthetic model (orange) and fine-tuned model (blue) on 79 real-world ECG images containing artefact. TPR = True positive rate, FPR = False positive rate.

## Discussion

In the present study, we utilised a publicly available signal-based dataset to create GenECG – a high-fidelity, synthetic image-based ECG dataset comprising 21,799 ECGs with artefacts encountered in routine clinical care paired with artefact-free images. Clinical Turing tests confirmed the ECGs were indistinguishable from real-world ECGs. Pre-existing image-based AI algorithms exhibited good performance levels on synthetic images without artefact, but poor performance on synthetic images with artefact. Importantly, existing algorithm accuracy on real-world ECG images could be improved following fine-tuning with GenECG data. These findings highlight the potential for synthetic ECG data to augment clinically useful image-based AI-ECG algorithm development.

Signal-based AI-ECG algorithms have previously demonstrated diagnostic capabilities comparable to experienced clinicians[21,22] and the ability to detect subtle ECG patterns, facilitating both improved screening and phenotyping of disease.[23] However, the requirement for signal data presents a barrier in many clinical areas where signal data are unavailable.[12] Such areas include low resource areas, pre-hospital settings and hospital environments where paper-based ECGs continue to be used. To address this issue, digitisation tools have been developed to derive signal data from paper-based ECGs.[24–26] Unfortunately, external validation of these tools is limited by a relative unavailability of image-based data, making it difficult to assess digitisation methods.[27] GenECG provides a benchmark dataset which could be used with PTB-XL signal data[7] to enable external validation of ECG digitisation tools. Alternatively, the development of image-based algorithms using GenECG could obviate the requirement to digitise paper-based ECGs altogether. Given the absence of a universal format for digitised ECG data storage and exchange,[28] this option would offer additional clinical utility.

In the present study, we used an existing signal-based dataset to create a large-scale image-based ECG dataset. In creating the images from the PTB-XL database, GenECG benefits from the PTB-XL dataset’s broad scope and variety.[7] Moreover, we have demonstrated the incorporation of artefacts into GenECG thereby ensuring that the dataset represents a real-world clinical dataset as closely as possible. Within our study, the series of ‘clinical Turing tests’ confirmed that the dataset that we have created is life-like with generated images incorporating the artefacts typically encountered with real-world ECGs. To our knowledge, GenECG is the first synthetic image-based ECG dataset to pass a Turing test.

The performance of pre-existing image-based algorithms on both real-world ECGs containing artefact and our GenECG data indicates poor generalisability of existing algorithms to real-world settings. In training and fine-tuning the model developed by Bridge *et al.*[18], we have demonstrated that utilising synthetic ECG data may overcome the limited efficacy of pre-existing image-based algorithms on real-world ECG images. Importantly, the performance of the fine-tuned model on real-world images has also demonstrated the potential for synthetic data to be used to improve the performance of image-based AI-ECG algorithms. Our synthetic ECG images provide a large data repository which can be used to facilitate the development of AI-ECG algorithms on images containing artefacts. This will ensure the translation of AI-ECG analysis from the research setting to the clinical workplace.

### Limitations

In this study, ECGs were reconstructed from clinically recorded signal data. Whilst the Turing tests confirmed the fidelity of the images created, whether fully synthetic ECGs images can be created is not yet known. A further limitation is that the ECGs created are of a single layout type. Whilst several major cardiac societies have advocated a standardised simultaneous lead format for paper-based ECGs,[13] ECG layouts can vary in clinical practice. It is therefore crucial to ensure that future image-based datasets capture this variability such that image-based AI algorithms can be applied to ECGs regardless of the ECG layout encountered.

## Conclusion

This study presents GenECG – a high-fidelity, synthetic image-based ECG dataset comprising 21,799 ECG images paired with artefact-free images and ECG signal data. Clinical Turing tests confirmed the fidelity of the synthetic images, demonstrating their potential to facilitate image-based AI ECG algorithm development. Our findings demonstrate the ability of synthetic data to enhance the performance of pre-existing image-based algorithms. GenECG will enable the development of image-based AI-ECG algorithms promising to bridge the gap between AI-ECG research and clinical practice. This will ensure that AI-ECG can be utilised in hospital settings, where paper-based ECGs continue to be used, and non-hospital settings including remote healthcare areas and pre-hospital settings.

## Funding

The study was funded by a University of Edinburgh Wellcome Trust iTPA award. The authors acknowledge the support of the British Heart Foundation Centre for Research Excellence Award III (RE/18/5/34216). The authors acknowledge the support of the British Heart Foundation (RG/20/4/34803). SEW is supported by the British Heart Foundation (FS/20/26/34952). IK is supported by the British Heart Foundation (FS/CRTF/21/24166).

## Disclosure of interest

No competing interests to declare.

## Author Contributions

NB, MOB and SEW conceived the concept. NB, MJ, MOB and SEW wrote the paper. MK, AG, SN and MON provided intellectual input into the draft of the manuscript. AB, DR, and RB created the synthetic ECG images. NB, IK, and KT designed and conducted the Turing tests. NB, KT, MJ, and MOB examined the performance of the synthetic ECG images on pre-existing image-based algorithms. NB and VV collected and organised the dataset. MOB and SEW are equally contributing senior authors. All authors read, edited, and approved the final version. The corresponding author attests that all listed authors meet authorship criteria and that no others meeting the criteria have been omitted. NB is a guarantor.

## Ethics statement

Ethical approval was obtained to conduct the clinical Turing tests described in the study by the King’s College London Biomedical Sciences, Dentistry, Medicine and Natural & Mathematical Sciences Research Ethics Panel (LRS-22/23-38259).

## Data availability statement

The ECG images described in the study were created from the PTB-XL database.[7] The ECG images will be used for a British Heart Foundation Data Science Centre open challenge (https://bhfdatasciencecentre.org/areas-unstructured-data-imaging-open-challenge/). Following this challenge, Dataset A and B will be made publicly available via a Creative Commons license.

## Patient and public involvement

The GenECG project was presented at a Guy’s and St Thomas’ NHS Foundation Trust cardiac support group meeting in November 2023. Members of the group felt that the development of a synthetic image-based ECG dataset would be an important advancement in enhancing ECG analysis.

## Data Availability

The ECG images described in the study were created from the PTB-XL database.[9] The ECG images will be used for a British Heart Foundation Data Science Centre open challenge (https://bhfdatasciencecentre.org/areas-unstructured-data-imaging-open-challenge/). Following this challenge, Dataset A and B will be made publicly available via a Creative Commons license.

## Notes

### Competing Interest Statement

The authors have declared no competing interest.

### Author Declarations

The Ethics committee/IRB of King's College London gave ethical approval for the clinical Turing tests performed as part of this study (LRS-22/23-38259).

### Summary of Updates

Manuscript abstract updated. Clinical implications section included. Link to the British Heart Foundation Data Science Centre open challenge updated.

## References

1. Gonzalesid A, Guruswamy G, Smith SR. Synthetic data in health care: A narrative review. PLOS Digital Health 2023;2:e0000082.

2. Attia ZI, Harmon DM, Behr ER et al. Application of artificial intelligence to the electrocardiogram. Eur Heart J 2021;42:4717–30.

3. Kwon J myoung, Jung MS, Kim KH et al. Artificial intelligence for detecting electrolyte imbalance using electrocardiography. Ann Noninvasive Electrocardiol 2021;26, DOI: 10.1111/ANEC.12839.

4. Attia ZI, Kapa S, Lopez-Jimenez F et al. Screening for cardiac contractile dysfunction using an artificial intelligence–enabled electrocardiogram. Nat Med 2019;25:70–4.

5. Attia ZI, Noseworthy PA, Lopez-Jimenez F et al. An artificial intelligence-enabled ECG algorithm for the identification of patients with atrial fibrillation during sinus rhythm: a retrospective analysis of outcome prediction. Lancet 2019;394:861–7.

6. Raghunath S, Ulloa Cerna AE, Jing L et al. Prediction of mortality from 12-lead electrocardiogram voltage data using a deep neural network. Nature Medicine 2020 26:6 2020;26:886–91.

7. Wagner P, Strodthoff N, Bousseljot RD et al. PTB-XL, a large publicly available electrocardiography dataset. Sci Data 2020;7:1–15.

8. Moody GB, Mark RG. The impact of the MIT-BIH arrhythmia database. IEEE Engineering in Medicine and Biology Magazine 2001;20:45–50.

9. Taddei A, Distante G, Emdin M et al. The European ST-T database: standard for evaluating systems for the analysis of ST-T changes in ambulatory electrocardiography. Eur Heart J 1992;13:1164–72.

10. Khan AH, Hussain M, Malik MK. ECG Images dataset of Cardiac and COVID-19 Patients. Data Brief 2021;34, DOI: 10.1016/J.DIB.2021.106762.

11. Sangha V, Mortazavi BJ, Haimovich AD et al. Automated multilabel diagnosis on electrocardiographic images and signals. Nat Commun 2022;13, DOI: 10.1038/S41467-022-29153-3.

12. Bodagh N, Ali O, Kotadia I et al. Feasibility of artificial intelligence-enhanced electrocardiogram (AI-ECG) analysis in the current clinical environment: An online survey. EP Europace 2023;25, DOI: 10.1093/EUROPACE/EUAD122.533.

13. AHA/ACCF/HRS Scientific Statement: Recommendations for the Standardization and Interpretation of the Electrocardiogram: Part I: The Electrocardiogram and Its Technology | Heart Rhythm Society.

14. Waveform Database Software Package (WFDB) for MATLAB and Octave v0.10.0.

15. Stucci — Blender Manual.

16. Chuquicusma MJM, Hussein S, Burt J et al. How to Fool Radiologists with Generative Adversarial Networks? A Visual Turing Test for Lung Cancer Diagnosis. Proceedings - International Symposium on Biomedical Imaging 2017;2018**-April**:240–4.

17. Veturi YA, Woof W, Lazebnik T et al. SynthEye: Investigating the Impact of Synthetic Data on Artificial Intelligence-assisted Gene Diagnosis of Inherited Retinal Disease. Ophthalmology Science 2023;3:100258.

18. Bridge J, Fu L, Lin W et al. Artificial intelligence to detect abnormal heart rhythm from scanned electrocardiogram tracings. J Arrhythm 2022;38:425–31.

19. Szegedy C, Vanhoucke V, Ioffe S et al. Rethinking the Inception Architecture for Computer Vision.

20. Lanczos C. An Iteration Method for the Solution of the Eigenvalue Problem of Linear Differential and Integral Operators 1. J Res Natl Bur Stand (1934) 1950;45.

21. Ribeiro AH, Ribeiro MH, Paixão GMM et al. Automatic diagnosis of the 12-lead ECG using a deep neural network. Nat Commun 2020;11, DOI: 10.1038/S41467-020-15432-4.

22. Hannun AY, Rajpurkar P, Haghpanahi M et al. Cardiologist-level arrhythmia detection and classification in ambulatory electrocardiograms using a deep neural network. Nat Med 2019;25:65–9.

23. Sau A, Ng FS. The emerging role of artificial intelligence enabled electrocardiograms in healthcare. BMJ Medicine 2023;2:e000193.

24. Ravichandran L, Harless C, Shah AJ et al. Novel Tool for Complete Digitization of Paper Electrocardiography Data. IEEE J Transl Eng Health Med 2013;1:1800107– 1800107.

25. Baydoun M, Safatly L, Hassan OKA et al. High Precision Digitization of Paper-Based ECG Records: A Step Toward Machine Learning. IEEE J Transl Eng Health Med 2019;7, DOI: 10.1109/JTEHM.2019.2949784.

26. Wu H, Patel KHK, Li X et al. A fully-automated paper ECG digitisation algorithm using deep learning. Scientific Reports 2022 12:1 2022;12:1–12.

27. Lence A, Extramiana F, Fall A et al. Automatic digitization of paper electrocardiograms – A systematic review. J Electrocardiol 2023;80:125–32.

28. Badilini F, Young B, Brown B et al. Archiving and exchange of digital ECGs: A review of existing data formats. J Electrocardiol 2018, DOI: 10.1016/j.jelectrocard.2018.07.028.

