## Supplementary Appendix for "GenECG: A synthetic image-based ECG dataset to augment artificial intelligence-enhanced algorithm development"

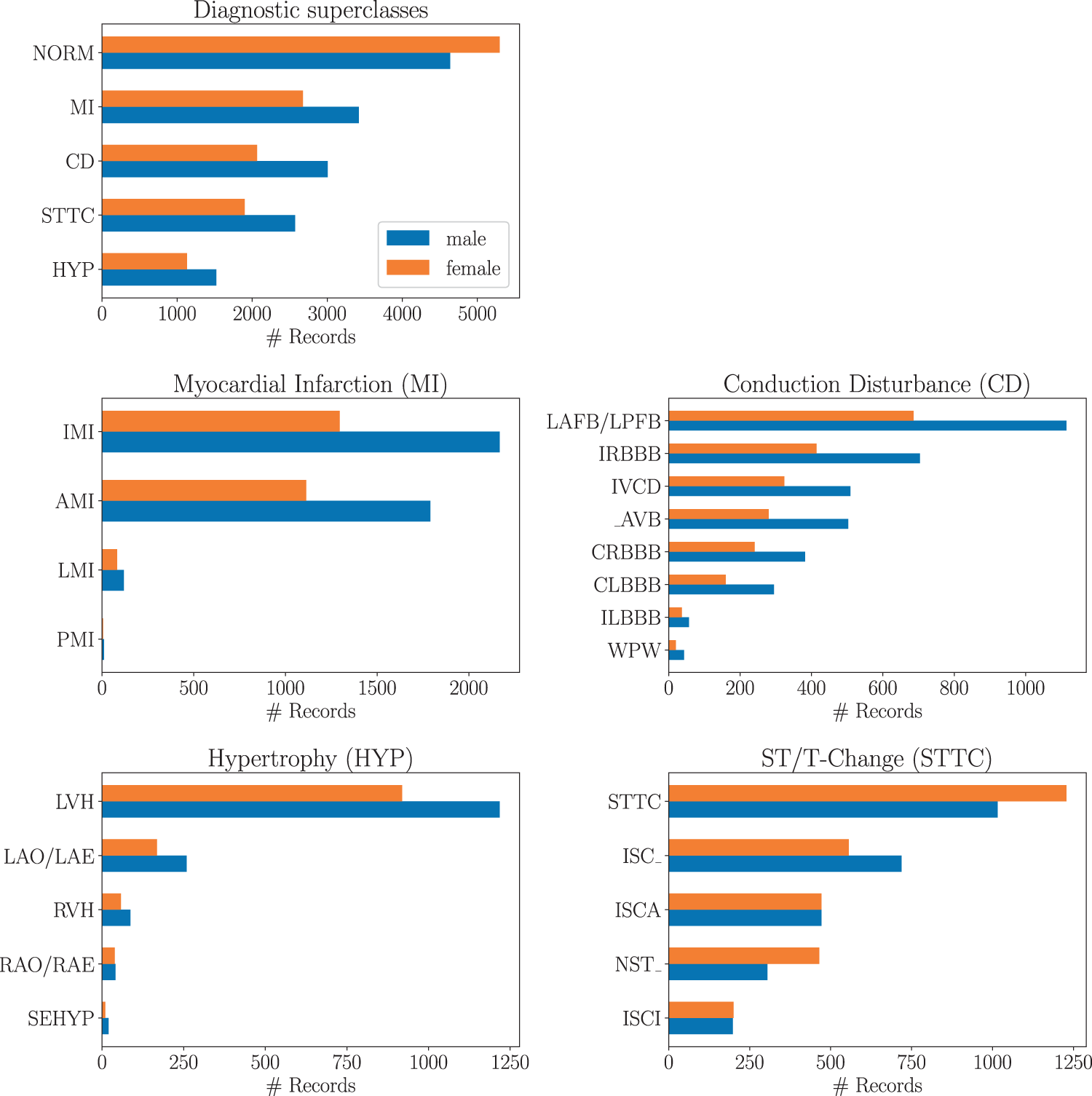

**Supplementary Figure S1.** PTB-XL dataset distribution of diagnostic subclasses for given diagnostic superclasses. Reproduced from Figure 5 of [1] under a Creative Commons Attribution 4.0 International License. See Supplementary Table S1 for SCP statement descriptions.

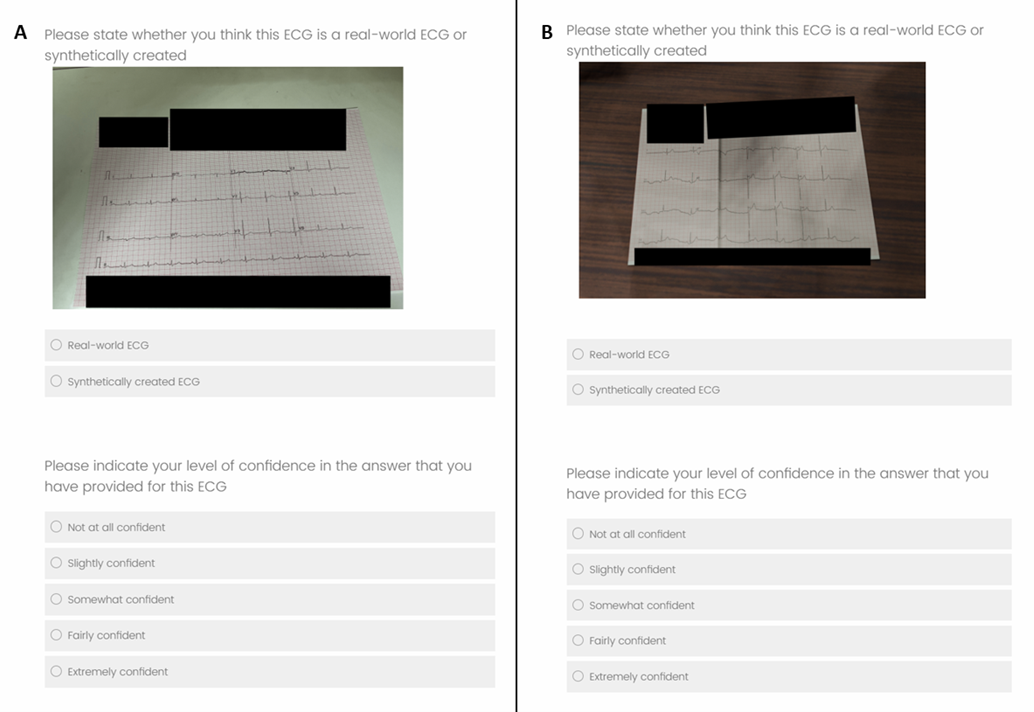

**Supplementary Figure S2.** Clinical Turing Tests. ECG images were individually displayed on pages and observers asked to select whether they thought each ECG image was a real-world ECG or synthetically created. For the second round and third rounds, observers were asked to indicate their levels of confidence in their answers using a five-point Likert scale. Figures (A) and (B) demonstrate two separate questions showing examples of (A) a real world-ECG and (B) a synthetically created ECG. Images were redacted in areas where text may appear.

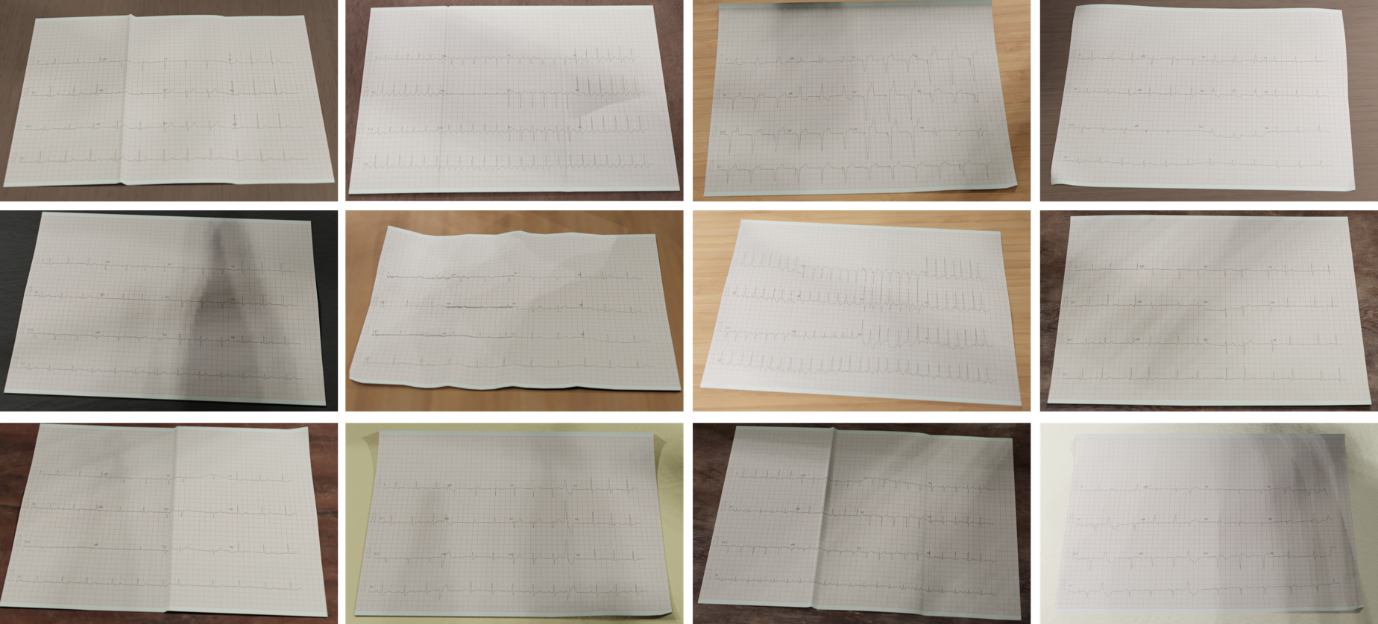
**Supplementary Figure S3.** Panel of 12 ECG images from “Dataset B: ECGs with artefact.”

|  | Acronym | SCP statement Description |  |
| --- | --- | --- | --- |
| Superclasses |  | NORM | Normal ECG |
|  |  | CD | Conduction Disturbance |
|  |  | MI | Myocardial Infarction |
|  |  | HYP | Hypertrophy |
|  |  | STTC | ST/T change |
| Subclasses | NORM | NORM | Normal ECG |
|  | CD | LAFB/LPFB | left anterior/left posterior fascicular block |
|  |  | IRBBB | incomplete right bundle branch block |
|  |  | ILBBB | incomplete left bundle branch block |
|  |  | CLBBB | complete left bundle branch block |
|  |  | CRBBB | complete right bundle branch block |
|  |  | _AVB | AV block |
|  |  | IVCB | non-specific intraventricular conduction disturbance (block) |
|  |  | WPW | Wolff-Parkinson-White syndrome |
|  | HYP | LVH | left ventricular hypertrophy |
|  |  | RHV | right ventricular hypertrophy |
|  |  | LAO/LAE | left atrial overload/enlargement |
|  |  | RAO/RAE | right atrial overload/enlargement |
|  |  | SEHYP | septal hypertrophy |
|  | MI | AMI | anterior myocardial infarction |
|  |  | IMI | inferior myocardial infarction |
|  |  | LMI | lateral myocardial infarction |
|  |  | PMI | posterior myocardial infarction |
|  | STTC | ISCA | ischemic in anterior leads |
|  |  | ISCI | ischemic in inferior leads |
|  |  | ISC_ | non-specific ischemic |
|  |  | STTC | ST-T changes |
|  |  | NST_ | non-specific ST changes |

**Supplementary Table S1.** PTB-XL dataset acronym descriptions for super- and subclasses. Reproduced from Table 5 of [1] under a Creative Commons Attribution 4.0 International License.

|  | Round One | Round Two | Round Three |
| --- | --- | --- | --- |
| Number of participants | 9 | 8 | 9 |
| Job role breakdown | 2 Consultants  5 Registrars  1 Senior House Officer  1 Nurse | 1 Consultant  3 Registrars  2 Senior House Officers  1 Physiologist  1 Nurse | 2 Consultants  2 Registrars  3 Senior House Officers  1 Physiologist  1 Advanced Clinical Practitioner |
| Total number of electrocardiograms correctly identified | 345/540 | 287/480 | 288/540 |
| Accuracy | 63.9% (95% CI 58.0%-69.8%) | 59.8% (95% CI 55.9%-63.7%) | 53.3% (95% CI 48.6%-58.1%) |
| Real world electrocardiograms correctly identified | 179/270 | 148/240 | 143/270 |
| True Recognition Rate | 66.3% (95% CI 60.4%-72.2%) | 61.7% (95% CI 55.3%-68.1%) | 53.0% (95% CI 48.7%-57.2%) |
| Synthetic electrocardiograms correctly identified | 166/270 | 139/240 | 145/270 |
| False Recognition Rate | 61.5% (95% CI 54.7%-68.3%) | 57.9% (95% CI 51.4%-64.5%) | 53.7% (95% CI 47.4%-60.0%) |
| Fleiss-Kappa Score | 0.045 (95% CI 0.003-0.087) | 0.003 (95% CI -0.044-0.051) | 0.049 (95% CI 0.007-0.092) |
| AUC-ROC score | Not performed | 0.565 (95% CI 0.514-0.616) | 0.480 (95% CI 0.432-0.529) |

**Supplementary Table S2.** Summarised results of iterative Turing tests. CI = Confidence interval. AUC-ROC = Area under the curve-receiver operating characteristic.

### References

1. Wagner P, Strodthoff N, Bousseljot RD *et al.* PTB-XL, a large publicly available electrocardiography dataset. *Sci Data* 2020;**7**:1–15.
